# Understanding Sociodemographic and Dietary Determinants of Cardiometabolic Risk: Cross-Sectional Evidence from the U.S. Healthy Eating Index to Inform Diet Quality Categories

**DOI:** 10.1101/2024.06.04.24308443

**Authors:** Elise Sheinberg, Laura A. Schmidt, Jerold R. Mande, Euridice Martinez-Steele, Deirdre K. Tobias, Cindy W. Leung

## Abstract

**Background:** Rising rates of diet-related chronic diseases and disparities in disease prevalence by sociodemographic factors highlight the need to improve the diet quality of Americans. Understanding how the Healthy Eating Index-2020 (HEI-2020) can be used as a measure of diet quality to create benchmarks for national nutrition monitoring can assist in surveillance and improve the design of nutrition programs and policies.

**Objective:** To examine the utility of the HEI-2020 to create benchmarks for national diet quality monitoring.

**Methods:** This serial cross-sectional study used data from the 2009-2018 National Health and Nutrition Examination Surveys (NHANES). Nationally representative data for 22,168 US adults (≥20 years) who completed two 24-hour recalls were analyzed. We derived HEI-2020 scores (0-100) from participants’ two 24-hour dietary recalls. Diet quality categories were established: high diet quality (>70-100), marginal (>60-70), low (>50-60), and very low (0-50).

**Results:** Only 13% of US adults had high diet quality while nearly two-thirds had low or very low diet quality. Diet quality was higher for older adults, female, “Other” race or ethnicity, born outside of the US, have higher education attainment, higher income, and food security. Compared to adults with high diet quality, adults with very low diet quality had lower intakes of unprocessed or minimally processed foods, fruits, vegetables, whole grains, and seafood and the highest intakes of ultra-processed foods, refined grains, and red and processed meats (all *P-trends <0.01*). Adults with very low diet quality were more likely to have elevated adiposity, lower HDL cholesterol, and higher triglycerides, fasting glucose, and hemoglobin A1c (all *P-trends* ≤*0.01*).

**Conclusions:** The HEI-2020 is a robust measure of diet quality that can be directly linked to biological measures associated with chronic disease risk. Using evidence-based HEI categories could allow policymakers, public health practitioners, and nutrition professionals to set benchmarks and nationwide targets for achieving improved diet quality.

## INTRODUCTION

Poor diet quality is a risk factor for chronic diseases, including obesity, diabetes, cardiovascular disease, stroke, and some cancers.^1–3^ Sociodemographic factors, including lower educational attainment, lower household income, minority racial and ethnic identity, and experiencing food insecurity, are associated with lower diet quality and higher chronic disease prevalence.^4–12^ Food insecurity, defined as “the limited or uncertain availability of nutritionally adequate and safe foods, or limited or uncertain ability to acquire acceptable foods in socially acceptable ways,”^13^ is associated with both poor diet quality and increased risk of chronic diseases.^14–16^ Concern about the relationship between food insecurity, hunger, and malnutrition led to the national monitoring of food insecurity since 1995.^17^ Validated food insecurity measurement tools can distinguish between individuals who have high food security, marginal food security, low food security, and very low food security.^13^ National trends and differences by sub-groups have informed public health policy and program development.

Nationally representative dietary intake is available through the National Health and Nutrition Examination Survey (NHANES), and can be used to assess alignment to the Dietary Guidelines for Americans (DGA), which provide diet patterns that prevent disease and promote health and well-being, with the Healthy Eating Index (HEI). According to the 2020-2025 DGA, a high-quality diet includes adequate vegetables, fruits, whole grains, low- and non-fat dairy, and low-fat protein, including beans and nuts, and limits consumption of added sugar, sodium, and saturated fat.^18^ The average HEI-2020 for adults was 56 out of 100 in 2023.^19^ Despite most Americans’ poor adherence to the DGA and rising rates of diet-related chronic diseases, diet quality categories that are associated with disease risk have not been established and used for national monitoring, as has been done for food insecurity.

There has been growing interest in the relationship between food security and diet quality, as well as a recognition of the need to address diet quality to reduce chronic diseases, constrain healthcare spending, and improve quality of life for all Americans.^20–22^ This has included bipartisan calls for policy action to monitor diet quality and consider the nutritional impact of federal nutrition programs, like the Supplemental Nutrition Assistance Program (SNAP).^23–25^ Population-based analyses of HEI have typically presented score means, and evidence to support the utility of specific thresholds (e.g. high vs. low diet quality) of risk for nutrition-related disease and mortality is warranted to understand if categories using the HEI can be used to monitor diet quality trends, similar to how food insecurity is monitored.^26–28^

Using nationally representative data, we created four diet quality categories of HEI-2020 scores: high (≥70), marginal (>60-70, low (>50-60), and very low (≤50) that were predictive of sociodemographic disadvantage and biomarkers associated with cardiometabolic disease risk and aligned with existing research.^3,29,30^ Establishing objective, evidence-based diet quality categories using the HEI can generate timely and critical data for policymakers, public health practitioners, and nutrition professionals to tackle the systemic drivers of poor diet quality and can promote long-term health for all Americans.

## METHODS

### Study Design and Population

We conducted our analyses using data from the 2009-2018 NHANES. NHANES is an ongoing multistage, nationally representative survey conducted by the National Center for Health Statistics (NCHS). In this study, the analytic sample included 22,168 adults ≥20 years with two valid 24-hour recalls (**Supplemental Figure 1**). Data collection was approved by NCHS ethics review board and all participants provided written consent.

### Dietary Assessment and the HEI-2020 scoring

Dietary intake was assessed via two 24-hour dietary recalls administered by trained interviewers using the USDA Automated Multi-Pass Method.^31^ One 24-hour recall is collected in-person, and a second recall is completed by telephone 3-10 days later. Methods and validation studies for conducting 24-hour recalls have been previously published.^32^ Data from the two 24-hour recalls were converted into standard food groups and food pattern components using the Food Patterns Equivalents Database (FPED) to calculate individual HEI-2020 scores.

Participants’ HEI-2020 scores were calculated using the simple HEI scoring algorithm method and averaged across the two days of intake (i.e., observed HEI). We also used the National Cancer Institute (NCI) SAS Macros for the Bivariate Method (version 2.1) in order to account for the within-person variability of dietary intake and to estimate predicted HEI scores (i.e., usual HEI).^33^

### HEI cut-points

The determination of the cut-points used to define categories of increasing diet quality was informed on multiple criteria. First, we comprehensively reviewed prior findings from prospective observational studies that examined HEI scores in relation to major chronic disease outcomes (**Supplemental Table 1**).^2^ Most studies derived population-specific quintiles of HEI scores as the exposure, ^34^ and individuals in the highest HEI quintile consistently had lower risks of cardiovascular disease, cancer, type 2 diabetes, and all-cause mortality compared to individuals in the lowest quintile.^2,35^ Second, we examined the distributions of observed and usual HEI scores in NHANES to understand the extent to which US adults aligned with the cut points in the studies above (**Supplemental Figure 2**). Finally, we considered NCI’s suggested HEI cut-points and corresponding grade.^36^ Collectively, this evidence informed the following category definitions: high diet quality (>70-100), marginal diet quality (>60-70), low diet quality (>50-60), and very low diet quality (0-50). High diet quality indicates a favorable adherence to the DGA and where the greatest risk reduction of morbidity and mortality from diet-related chronic diseases has been observed.^2,29,37,38^. In contrast, very low diet quality indicates poor adherence to the DGA and is associated with the highest risks of morbidity and mortality from diet-related chronic diseases. Marginal and low diet quality are intermediate risk levels.

### Other Dietary Components

All dietary variables were derived from the average of two days of dietary recalls and standardized to 1,000 kcal. Other dietary factors in the current analysis included total energy intake (kcal/day). whole fruits (cup equivalents/day), vegetables excluding white potatoes (cup equivalents), refined grains (ounce equivalents/day), whole grains (ounce equivalents/day), red and processed meats (ounce equivalents/day), and seafood (ounce equivalents/day). Nutrients of interest included saturated fat (g/day), dietary fiber (g/day), sodium (mg/day), potassium (mg/day), calcium (mg/day), and added sugar (tsp equivalents/day). We also applied the NOVA classification to derive intakes of unprocessed and minimally processed foods and ultra-processed foods (% energy/day).^39,40^

### Adiposity and cardiometabolic markers

Height, weight, and waist circumference (WC) were measured by trained personnel in the Mobile Examination Center.^41^ Body mass index was calculated as weight (kg) over height squared (m^2^). Obesity was defined as a BMI ≥30 kg/m^2^.^42^ High waist circumference was defined as >88 centimeters for females and >102 centimeters for males.^43^

Cardiometabolic biomarkers in this analysis include blood levels of high-density lipoprotein cholesterol (HDL-c; mg/dL), low-density lipoprotein cholesterol (LDL-c; mg/dL), triglycerides (mg/dL), and glucose (mg/dL). Low HDL cholesterol was defined as <40 mg/dL and elevated LDL cholesterol was defined as ≥100 mg/dL.^44^ Elevated triglycerides were defined as ≥150 mg/dL.^45^ Elevated fasting glucose was defined as >100 mg/dL.^46^

### Sociodemographic Characteristics

Characteristics that have been associated with diet quality in previous research were included as covariates.^47–49^ Self-reported sociodemographic variables were ascertained at patient visits, including age, sex, race and ethnicity, place of birth, educational attainment, employment status, marital status, family income to poverty ratio. Household food security status was derived using the 18-item U.S. Food Security Survey Module.^50^

### Statistical Analysis

Complex two-day dietary sampling weights were adjusted to reflect the 10-year analytic period and applied to all analyses to make nationally representative inferences. We first calculated the proportions US adults for each of our new HEI-based diet-quality categories cumulatively and separately for each NHANES 2-year cycle. We evaluated whether diet quality categories differed over time and by sociodemographic characteristics using the Satterwaite-adjusted Chi-Square test. We generated radar plots to visualize the contribution of HEI-components across diet quality categories, as a percentage of the maximum possible score for that component. For other individual dietary factors and cardiometabolic biomarkers, we did complete case analysis and estimated their least squared means across diet quality categories using multivariable linear regression models adjusted for all sociodemographic characteristics to improve precision. We estimate the predicted probabilities of the clinically relevant cardiometabolic biomarkers in each diet quality category with logistic regression models, adjusting for all sociodemographic characteristics. In sensitivity analyses, statistical tests were repeated using diet quality categories derived from usual HEI scores. Statistical analyses were performed with SAS 9.4 (SAS Institute Inc., Cary, NC).

## RESULTS

Between 2009 and 2018, 12.7% of US adults had high diet quality (HEI ≥70), 19.9% had marginal diet quality (HEI 60-<70), 26.9% had low diet quality (HEI 50-<60), and 40.5% had very low diet quality (HEI ≤50) (**Figure 1**). When usual dietary intakes were used, only 0.8% of US adults had high diet quality, 12.9% had marginal diet quality, 41.3% had low diet quality, and 45.0% of adults had very low diet quality. There were no significant differences over the 10-year NHANES cycles in the study period (**Supplemental Figure 3**, *P>0.05)*

**Figure 1:**
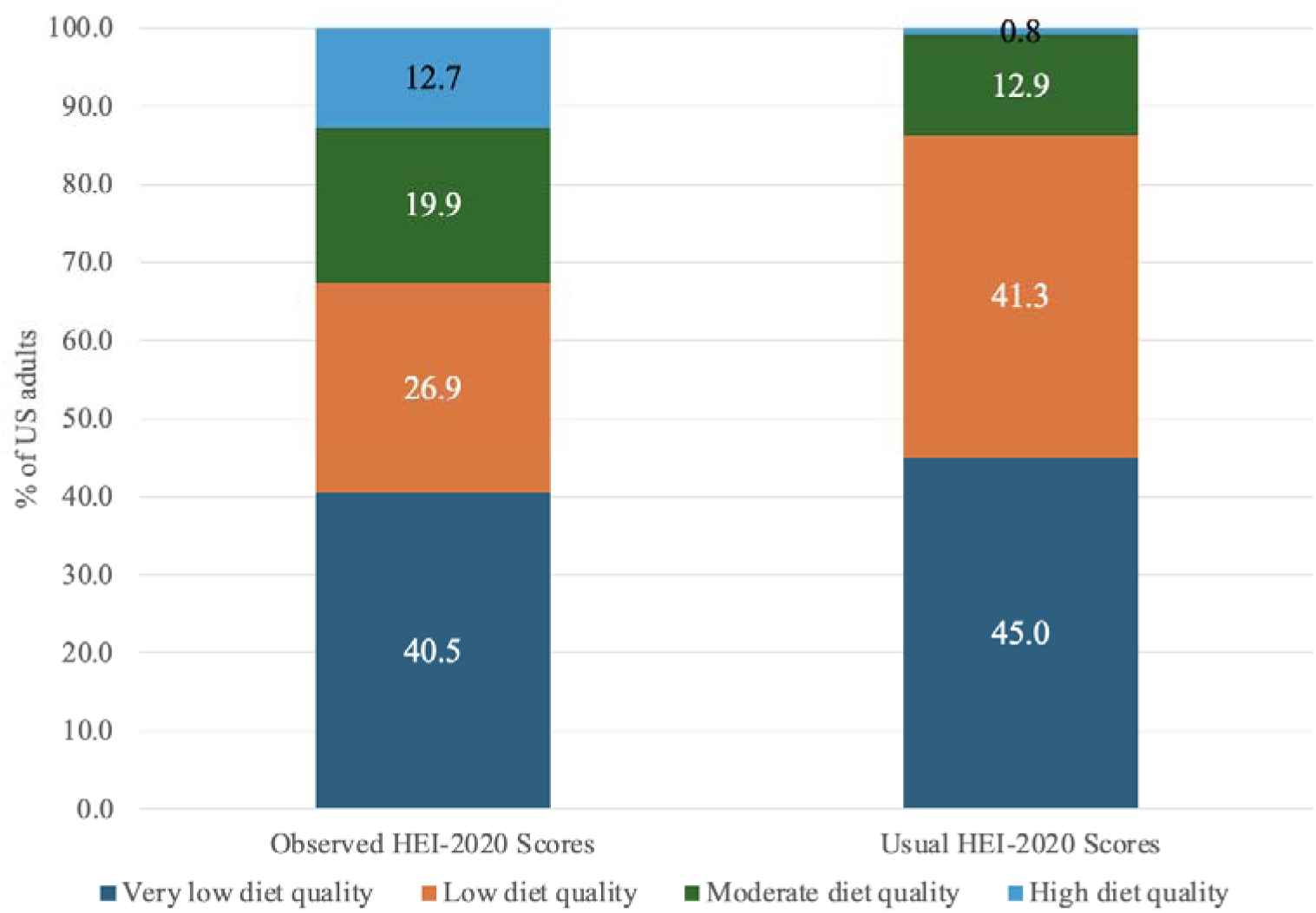
**Proportion of diet quality categories using observed and usual HEI-2020 scores**

There were statistically significant differences in diet quality categories by all sociodemographic characteristics except employment status (**Table 1**, *P-linear trend <0.01*). Adults ≥51 years were more likely to have high diet quality (16.1%) than adults ≤30 years (8.0%). Females were more likely to have high diet quality than males (14.9% compared with 10.4%, respectively). By race and ethnicity, 8.0% of non-Hispanic Black adults had high diet quality compared to 11.0% of Hispanic adults, 13.1% of non-Hispanic white adults, and 19.4% of adults in another racial or ethnic group (i.e., “Other”). Adults born outside of the US were more likely to have high diet quality (19.2%) than adults born in the US (11.5%). Adults with some college education or more were more likely to have high diet quality (15.5%) than those with a high school education or less (8.0%). Married or partnered adults (13.7%) were more likely to have high diet quality than single adults (11.1%). Adults with household incomes >200% FPL were more likely to have high diet quality (14.6%) than adults with household incomes ≤200% FPL (9.0%). Adults with high or marginal food security were more likely to have high diet quality (13.9%) than adults with low or very low food security (6.3%).

**Table 1:** Sociodemographic characteristics of US adults (. ≥**20 years) stratified by diet quality categories using observed intake, NHANES 2009-2018^c^**

|  | Overall<br>N (weighted %) | Diet quality category |  |  |  |
| --- | --- | --- | --- | --- | --- |
|  |  | Very low<br>(HEI 0-50)<br>N (weighted %) | Low<br>(HEI >50-60)<br>N (weighted %) | Marginal<br>(HEI >60-70)<br>N (weighted %) | High<br>(HEI >70-100)<br>N (weighted %) |
| Age |  |  |  |  |  |
| $\leq 30$ years | 3917 (20.3) | 2040 (51.6) | 994 (25.3) | 574 (15.1) | 309 (8.0) |
| 31-50 years | 7396 (35.3) | 3229 (42.8) | 1949 (27.2) | 1392 (18.7) | 826 (11.2) |
| $\geq 51$ years | 10855 (44.4) | 3640 (33.5) | 2945 (27.3) | 2458 (23.1) | 1812 (16.1) |
| Sex |  |  |  |  |  |
| Male | 10548 (48.1) | 4616 (44.6) | 2830 (27.1) | 1943 (17.9) | 1159 (10.4) |
| Female | 11620 (51.9) | 4293 (36.7) | 3058 (26.7) | 2481 (21.8) | 1788 (14.9) |
| Race and ethnicity |  |  |  |  |  |
| Hispanic | 5331 (14.6) | 1959 (40.9) | 1488 (27.3) | 1183 (20.8) | 701 (11.0) |
| Non-Hispanic Black | 4849 (11.3) | 2176 (47.9) | 1331 (27.5) | 847 (16.5) | 495 (8.0) |
| Non-Hispanic White | 9219 (65.7) | 3939 (40.0) | 2376 (26.9) | 1732 (20.0) | 1172 (13.1) |
| Other<sup>a</sup> | 2769 (8.4) | 835 (33.0) | 693 (24.9) | 662 (22.7) | 579 (19.4) |
| Birthplace |  |  |  |  |  |
| United States | 16599 (83.8) | 7458 (42.7) | 4407 (27.0) | 2955 (18.8) | 1779 (11.5) |
| Outside of the United States | 5569 (16.2) | 1451 (28.8) | 1481 (26.4) | 1469 (25.5) | 1168 (19.2) |
| Educational attainment |  |  |  |  |  |
| High school education or less | 9723 (36.4) | 4472 (49.4) | 2615 (26.2) | 1723 (16.4) | 913 (8.0) |
| Any college or college graduate | 12445 (63.6) | 4437 (35.4) | 3273 (27.3) | 2701 (21.9) | 2034 (15.5) |
| Employment status |  |  |  |  |  |
| Employed | 12172 (61.2) | 5007 (41.3) | 3225 (26.6) | 2382 (19.9) | 1558 (12.2) |
| Not employed | 9996 (38.8) | 3902 (39.2) | 2663 (27.3) | 2042 (20.0) | 1389 (13.6) |
| Marital status |  |  |  |  |  |
| Married or living with partner | 13214 (63.1) | 5000 (38.3) | 3528 (26.9) | 2786 (21.1) | 1900 (13.7) |
| Single | 8954 (36.9) | 3909 (44.1) | 2360 (26.8) | 1638 (17.9) | 1047 (11.1) |
| Household income |  |  |  |  |  |
| ≤200% federal poverty level | 9690 (33.0) | 4504 (48.7) | 2533 (25.7) | 1690 (16.6) | 963 (9.0) |
| >200% federal poverty level | 12478 (67.0) | 4405 (36.4) | 3355 (27.5) | 2734 (21.6) | 1984 (14.6) |
| Household food security |  | 4405 (36.4) | 3355 (27.5) | 2734 (21.6) | 1984 (14.6) |
| High/marginal food security | 17534 (84.2) | 6571 (37.8) | 4745 (27.4) | 3642 (20.9) | 2576 (13.9) |
| Low/very low food security | 4634 (15.8) | 2338 (54.9) | 1143 (24.1) | 782 (14.6) | 371 (6.3) |

<sup>a</sup> Other included American Indian or Alaskan Native, Asian, Native Hawaiian or Pacific Islander, and Other
<sup>b</sup> Household food security assessed using the U.S. Household Food Security Survey Module

A radar plot showing the proportion of the possible points for each HEI component for each diet quality category is shown in **Figure 2**. Adults with high diet quality scored at least half of the possible points for all HEI components. Adults with very low diet quality scored less than half of the possible points for nine components. The greatest differences in HEI-2020 component scores between adults with high versus very low diet quality were for whole grains (very low diet quality scored 78.4% lower than high diet quality), whole fruit (very low diet quality scored 72.5% lower than high diet quality), total fruit (very low diet quality scored 70.9% lower than high diet quality), greens and beans (very low diet quality scored 69.1% lower than high diet quality), seafood and plant protein (very low diet quality scored 58.9% lower than high diet quality) and fatty acids (very low diet quality scored 55.4% lower than high diet quality).

**Figure 2:**
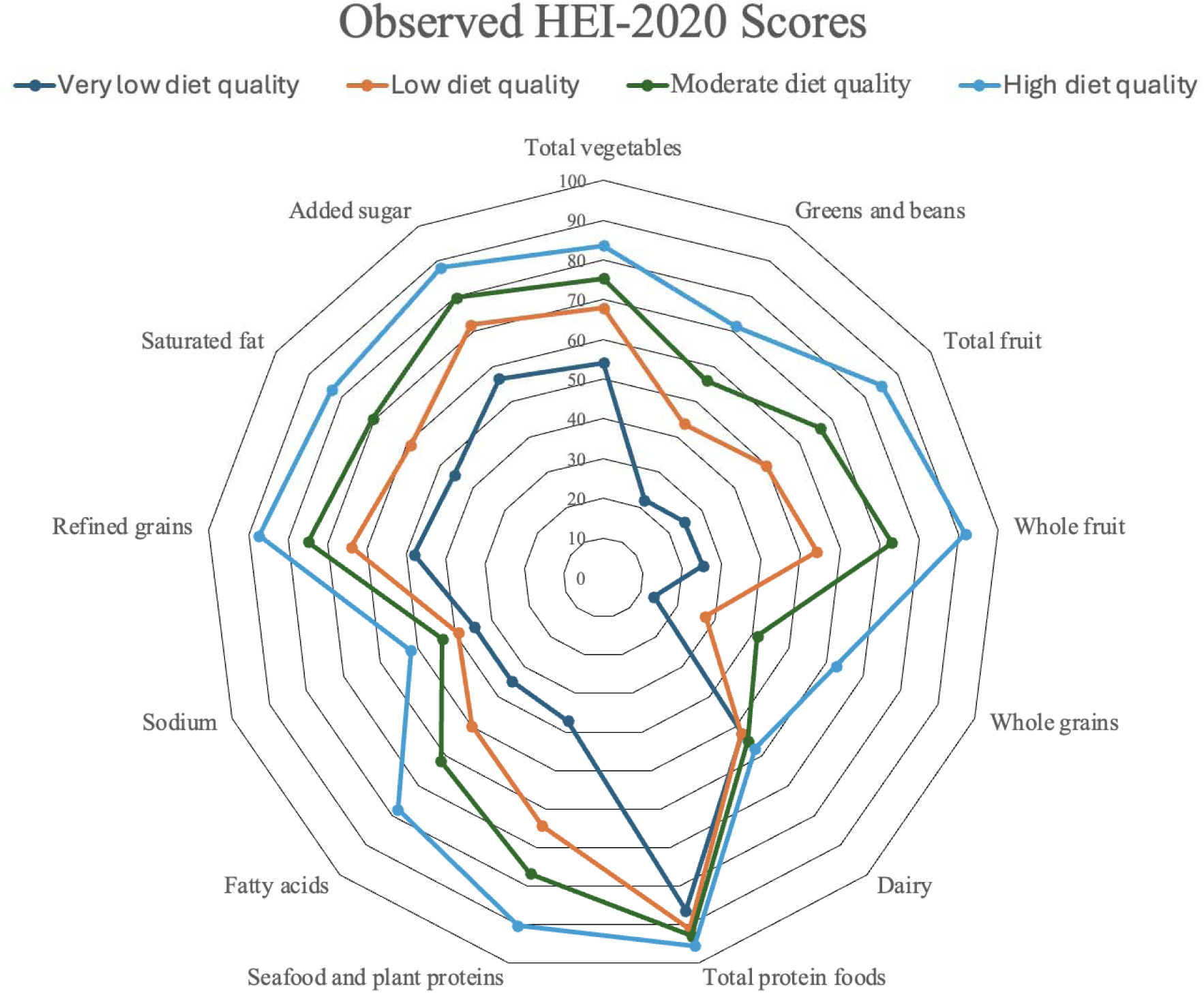
**HEI-2020 component scores by diet quality categories, 2009-2018 NHANES**

Multivariate-adjusted mean intakes of foods and nutrients of interest across diet quality categories are shown in **Table 2**. Mean total energy intake was lowest among adults with high diet quality (1976 kcal/day) and increased across diet quality categories (marginal: 2031 kcal/day, low: 2102 kcal/day; and very low: 2106 kcal/day) (*P-linear trend<0.01*). Adults with high diet quality had the highest intakes of unprocessed or minimally processed foods (41.4% energy) and adults with very low diet quality had the lowest intakes (25.8% energy) – a difference of +15.6% (*P-linear trend<0.01*). Similarly, adults with high diet quality had the lowest intakes of ultra-processed foods (42.5% energy) while adults with very low diet quality had the highest intakes (61.7% energy)- a difference of −19.2% (*P-linear trend<0.01*).

**Table 2:** Multivariate-adjusted observed daily intakes of total energy, NOVA food groups, foods, and nutrients of interest stratified by diet quality categories (using observed HEI-2020 scores), NHANES 2009-2018.

|  | Overall<br>N | Diet quality category |  |  |  | *P-linear*<br>*trend* |
| --- | --- | --- | --- | --- | --- | --- |
|  |  | Very low<sup>a</sup><br>Mean | Low<sup>a</sup><br>Mean | Marginal<sup>a</sup><br>Mean | High<sup>a</sup><br>Mean |  |
| Total energy (kcal/day) | 2085 | 2106<sup>b</sup> | 2102<sup>b</sup> | 2031<sup>b</sup> | 1976 | <0.01 |
| Nova 1-2, Unprocessed/ minimally<br>processed foods (% energy/day) | 30.1 | 25.8<sup>b</sup> | 32.4<sup>b</sup> | 36.3<sup>b</sup> | 41.4 | <0.01 |
| NOVA 4, Ultra-processed foods (%<br>energy/day) | 55.2 | 61.7<sup>b</sup> | 53.0<sup>b</sup> | 48.5<sup>b</sup> | 42.5 | <0.01 |
| Foods (cup or oz eq per 1000 kcal) |  |  |  |  |  |  |
| Fruits, excluding juice (cup eq/day) | 0.7 | 0.3<sup>b</sup> | 0.7<sup>b</sup> | 1.1<sup>b</sup> | 1.6 | <0.01 |
| Vegetables, excluding white potatoes (cup<br>eq/day) | 0.6 | 0.5<sup>b</sup> | 0.6<sup>b</sup> | 0.7<sup>b</sup> | 1.0 | <0.01 |
| Refined grains (oz eq/day) | 5.5 | 6.8<sup>b</sup> | 5.7<sup>b</sup> | 4.7<sup>b</sup> | 3.7 | <0.01 |
| Whole grains (oz eq/day) | 0.9 | 0.4<sup>b</sup> | 0.9<sup>b</sup> | 1.3<sup>b</sup> | 2.1 | <0.01 |
| Red and processed meat (oz eq/day) | 2.6 | 2.9<sup>b</sup> | 2.5<sup>b</sup> | 2.2<sup>b</sup> | 1.6 | <0.01 |
| Seafood (oz eq/day) | 0.6 | 0.4<sup>b</sup> | 0.7<sup>b</sup> | 1.4 | 1.7 | <0.01 |
| Nutrients per 1,000 calories |  |  |  |  |  |  |
| Saturated fat (% energy/day) | 26.3 | 29.1<sup>b</sup> | 25.8<sup>b</sup> | 22.1<sup>b</sup> | 18.5 | <0.01 |
| Dietary fiber (g/day) | 17.3 | 13.4<sup>b</sup> | 17.4<sup>b</sup> | 20.1<sup>b</sup> | 25.5 | <0.01 |
| Sodium (mg/day) | 3481 | 3663<sup>b</sup> | 3528<sup>b</sup> | 3307<sup>b</sup> | 3007 | <0.01 |
| Potassium (mg/day) | 2670 | 2249<sup>b</sup> | 2676<sup>b</sup> | 2907<sup>b</sup> | 3221 | <0.01 |
| Calcium (mg/day) | 967 | 908<sup>b</sup> | 957<sup>b</sup> | 982<sup>b</sup> | 1049 | <0.01 |
| Added sugar (% energy/day) | 16.4 | 20.3<sup>b</sup> | 15.6<sup>b</sup> | 13.2<sup>b</sup> | 10.8 | <0.01 |

<sup>a</sup> Values adjusted for age, sex, race and ethnicity, birthplace, educational attainment, employment status, marital status, household income, and household food security
<sup>b</sup> Significant difference compared to the high diet quality category

Adults with high diet quality consumed significantly more fruits, vegetables, whole grains, and seafood, and less refined grains and red and processed meats when compared to adults with marginal, low, and very low diet quality (all *P-linear trends<0.01)* (**Table 2**). Adults with high diet quality also consumed lower amounts of saturated fat, sodium, and added sugars and higher amounts of dietary fiber, potassium, and calcium than adults with marginal, low, and very low diet quality (all *P-linear trends<0.01*). Nearly all comparisons between the high diet quality group with marginal, low, and very low diet quality were statistically significant, except for seafood consumption between high and marginal diet quality groups. Sensitivity analyses examining multivariate-adjusted means of usual intakes showed identical patterns (**Supplemental Table 2**).

There were differences in BMI and obesity prevalence across diet quality categories (**Table 3)**. After adjusting for sociodemographic factors, the mean BMI for adults with high diet quality was lower (27.4 kg/m^2^) compared to adults with marginal (28.2 kg/m^2^ ), low (29.2 kg/m^2^), and very low diet quality (29.8 kg/m^2^) (*P-linear trend<0.01*). Relatedly, 27.3% of adults with high diet quality had obesity compared to 45.7% of adults with very low diet quality – a difference of −18.4% (*P-trend<0.01*). Adults with high diet quality had a mean waist circumference of 94.0 cm (49.3% with elevated waist circumference), compared to a mean waist circumference of 100.4 cm for adults with very low diet quality (61.5% elevated waist circumference) (*P-linear trends<0.01*).

**Table 3:** Multivariate-adjusted adiposity and cardiometabolic markers stratified by diet quality categories (observed HEI-2020 scores), NHANES 2009-2018.

|  | Overall | Diet quality category |  |  |  | *P-linear trend* |
| --- | --- | --- | --- | --- | --- | --- |
|  |  | Very Low<sup>a</sup> | Low<sup>a</sup> | Marginal<sup>a</sup> | High<sup>a</sup> |  |
| BMI (kg/m2)<sup>c</sup> | 29.3 | 29.8 (0.1)<sup>b</sup> | 29.2 (0.2)<sup>b</sup> | 28.2 (0.2)<sup>b</sup> | 27.4 (0.2) | <0.01 |
| % with obesity<sup>d</sup> | 38.8 | 45.7 (8.4)<sup>b</sup> | 41.5 (7.9)<sup>b</sup> | 33.8 (7.2)<sup>b</sup> | 27.3(6.1) | <0.01 |
| Waist circumference (cm)<sup>c</sup> | 99.7 | 100.4 (0.3)<sup>b</sup> | 98.6 (0.3)<sup>b</sup> | 96.5 (0.4)<sup>b</sup> | 94.0 (0.5) | <0.01 |
| % with high WC<sup>d</sup> | 57.8 | 61.5 (16.4)<sup>b</sup> | 60.2 (16.4)<sup>b</sup> | 55.9 (16.6) | 49.3 (16.1) | <0.01 |
| HDL cholesterol (mg/dL)<sup>c</sup> | 53.6 | 50.7 (0.3)<sup>b</sup> | 52.9 (0.4)<sup>b</sup> | 54.1 (0.4)<sup>b</sup> | 56.6 (0.5) | <0.01 |
| % <40 mg/dL<sup>d</sup> | 18.6 | 24.1 (12.0)<sup>b</sup> | 19.1 (10.4) | 15.9(9.1) | 13.5 (8.0) | <0.01 |
| LDL cholesterol (mg/dL)<sup>c</sup> | 113.4 | 112.6 (0.7)<sup>b</sup> | 111.6 (1.0)<sup>b</sup> | 111.9 (1.0)<sup>b</sup> | 107.8 (1.4) | 0.01 |
| % ≥100 mg/dL<sup>d</sup> | 62.8 | 63.1 (9.7) | 63.1 (9.1) | 65.3 (8.5) | 61.6 (8.5) | 0.14 |
| Triglycerides (mg/dL)<sup>c</sup> | 120.4 | 123.7 (2.4)<sup>b</sup> | 120.0 (2.0)<sup>b</sup> | 118.4 (2.6)<sup>b</sup> | 103.6 (2.6) | <0.01 |
| % ≥150 mg/dL<sup>d</sup> | 23.2 | 25.7 (7.6)<sup>b</sup> | 25.8 (7.3)<sup>b</sup> | 23.9 (6.6) | 18.6 (5.2) | <0.01 |
| Fasting glucose (mg/dL)<sup>c</sup> | 106.8 | 107.1 (0.5)<sup>b</sup> | 105.4 (0.6)<sup>b</sup> | 106.1 (0.9)<sup>b</sup> | 101.7 (0.8) | <0.01 |
| % >100 mg/dL<sup>d</sup> | 42.9 | 47.7 (19.6)<sup>b</sup> | 48.6 (19.1)<sup>b</sup> | 44.6 (18.0) | 43.0 (17.3) | <0.01 |

<sup>a</sup> Values adjusted for age, sex, race and ethnicity, birthplace, educational attainment, employment status, marital status, household income, and household food security
<sup>b</sup> Significant difference compared to the high diet quality category
<sup>c</sup> Mean (Standard Error)
<sup>d</sup> Mean (Standard Deviation)

Cardiometabolic biomarkers predictive of diet-related disease also differed across diet quality categories (**Table 3**). Mean HDL cholesterol was higher among adults with high diet quality (56.6 mg/dL) and decreased across diet quality categories (marginal: 54.1 mg/dL; low: 52.9 mg/dL; very low: 50.7 mg/dL) (*P-trend<0.01*). Clinically low HDL cholesterol was observed in 13.5% of adults with high diet quality compared to 24.1% of adults with very low diet quality – a difference of 10.6% (*P-linear trend<0.01*). Similarly, adults with high diet quality had lower mean LDL cholesterol levels (107.8 mg/dL) compared to adults with marginal (111.9 mg/dL), low (111.6 mg/dL), and very low diet quality (112.6 mg/dL) (*P-linear trend=0.01*). There was no difference in the proportion of adults with elevated LDL cholesterol by diet quality category (*P-linear trend=0.14*). Adults with high diet quality had a mean triglyceride level of 103.6 mg/dL and 18.6% had clinically elevated triglycerides; adults with very low diet quality had a mean triglyceride level of 123.7 mg/dL, and 25.7% had clinically elevated triglycerides (*P-linear trends<0.01*). Adults with high diet quality had lower fasting glucose (101.7 mg/dL) compared to adults with very low diet quality (mean fasting glucose 107.1 mg/dL) (*P-linear trends<0.01*). Approximately 43% of adults with high diet quality had clinically elevated fasting glucose compared to 44.6% of adults with marginal, 48.6% with low, and 47.7% with very low diet quality (*P-linear trend=<0.01*). In sensitivity analyses, multivariate-adjusted differences in adiposity and cardiometabolic biomarkers by HEI categories using usual intake revealed consistent patterns (**Supplemental Table 3**)

## DISCUSSION

In this nationally representative ten-year study, only 13% of adults had high diet quality, and most (67%) had low or very low diet quality. These proportions were more magnified when considering usual HEI scores. This indicates that few Americans consume a diet aligned with the DGA at levels that protect against diet-related chronic disease morbidity and mortality, with notable disparities across sociodemographic factors. There were also no observable improvements in the proportion of adults achieving high diet quality over time.^24^

We noted disparities in diet quality categories by key sociodemographic characteristics. Non-Hispanic Black race/ethnicity, having a high school education or less, low household income, and food insecurity were all significantly associated with higher prevalence of very low diet quality. Although these are cross-sectional data, prior evidence indicates structural factors contribute to disparities in diet quality including racism, limited educational opportunities, inequitable food access, a lack of proper training, resources, or enough time to prepare healthy meals.^10,51–53^

In this study, more healthful diet quality categories were associated with a healthy weight status, lipid profile, and glycemic control in a dose-response manner. This is consistent with meta-analyses of prospective observational studies, which found that individuals who score at the top quintile of diet quality for their population have lower risks of all-cause mortality, cardiovascular disease, cancer, and type 2 diabetes compared to individuals who score in the bottom quintile.^2,38^ This indicates that the proposed HEI cut-points reflect the underlying cardiometabolic risk profile of US adults.

Significant differences in all foods, food groups, and nutrients of interest were observed across diet quality categories. Shifting to a higher diet quality category can be achieved through incremental dietary improvements. For example, the difference between high and marginal diet quality categories could be translated to a daily 6% kcal reduction from ultra-processed foods, 0.5 cups more fruits, 0.3 cups more vegetables, 0.8 oz equivalents more whole grains, and 0.3 oz equivalents more seafood. Similarly, the difference between marginal and low diet quality categories was, on average, a daily 5.5% kcal reduction from ultra-processed foods, 0.4 cups more fruits, 0.1 cups more vegetables 0.4 oz equivalents more whole grains and 0.7 oz equivalents more seafood. Finally, the difference between low and very low diet quality was on average an 8.7% reduction of energy/day from ultra-processed foods, 0.4 cups/day more fruit, 0.1 cups/day more vegetables, 0.5 oz equivalents/day more whole grains, and 0.3-ounce equivalents/day more seafood.

There have been calls to supplement food security surveillance measures with diet quality measures.^22,23,25,54^ This study shows that defining cut-points of HEI-2020 scores adequately and comprehensively captures underlying differences in key dietary, biological, and social risk factors implicated in diet-related diseases and could be used to efficiently assess the diet quality of the population. Nutrition interventions that work with individuals who have risk factors for very low diet quality often monitor food insecurity status of participants, and using evidence-based diet quality targets may enhance efforts to monitor and improve diet quality. This can reduce disparities in diet-related chronic diseases by raising participants into higher diet quality categories, in combination with other program evaluations and benchmarks.

Strengths of this study include our use of data from a large nationally representative sample of US adults spanning recent years. Second, we identified the cut points for HEI categories through multiple sources of health outcome evidence. Third, we leveraged the rich data sources available from NHANES, validated dietary recalls, physical measurements, and available biomarker data, to examine concurrent implications of the diet quality categories at the individual level. However, this study is limited by its cross-sectional nature; prospective follow-up is needed to capture clinical outcomes. The 24-hour recall is a validated approach but is subject to measurement error and recall bias. Cut-points could result in misclassification of individuals. Furthermore, we did not account for the presence of chronic diseases or use of medications, as the goal was not to show causal effects of diet quality categories and biomarker outcomes, but to characterize population-level differences across diet quality categories.

## Conclusion

Given that the HEI is already used to monitor the nutritional status of the US, using HEI cut-points to establish diet quality categories would be an efficient, evidence-based, and practical approach allow policy makers, public health practitioners and nutrition professionals to set benchmarks and targets for nutrition policies and interventions. Disaggregating continuous HEI scores into meaningful categories can better identify disparities in diet-related health outcomes and help tailor existing nutrition programs and policies to improve healthful food access, increase diet quality, and promote health for all Americans.

## Author Contributions

CWL LAS, JRM and DKT designed research; EMS provided essential materials; CWL analyzed data; ES and CWL wrote paper; ES had primary responsibility for the final content; all authors read and approved the final manuscript; all authors have read and approved the final manuscript.

## Data Availability

All data produced in the present study are available upon reasonable request to the authors

https://wwwn.cdc.gov/nchs/nhanes/Default.aspx

## REFERENCES

1. Micha R, Peñalvo JL, Cudhea F, Imamura F, Rehm CD, Mozaffarian D. Association Between Dietary Factors and Mortality From Heart Disease, Stroke, and Type 2 Diabetes in the United States. JAMA. 2017;317(9):912–924. doi:10.1001/jama.2017.0947

2. Morze J, Danielewicz A, Hoffmann G, Schwingshackl L. Diet Quality as Assessed by the Healthy Eating Index, Alternate Healthy Eating Index, Dietary Approaches to Stop Hypertension Score, and Health Outcomes: A Second Update of a Systematic Review and Meta-Analysis of Cohort Studies. J Acad Nutr Diet. 2020;120(12):1998–2031.e15. doi:10.1016/j.jand.2020.08.076

3. Xu Z, Steffen LM, Selvin E, Rebholz CM. Diet quality, change in diet quality and risk of incident CVD and diabetes. Public Health Nutr. 2020;23(2):329–338. doi:10.1017/S136898001900212X

4. National Diabetes Statistics Report | Diabetes | CDC. November 20, 2023. Accessed November 24, 2023. https://www.cdc.gov/diabetes/data/statistics-report/index.html

5. Lopez-Neyman SM, Davis K, Zohoori N, Broughton KS, Moore CE, Miketinas D. Racial disparities and prevalence of cardiovascular disease risk factors, cardiometabolic risk factors, and cardiovascular health metrics among US adults: NHANES 2011–2018. Sci Rep. 2022;12(1):19475. doi:10.1038/s41598-022-21878-x

6. Schultz WM, Kelli HM, Lisko JC, et al. Socioeconomic Status and Cardiovascular Outcomes. Circulation. 2018;137(20):2166–2178. doi:10.1161/CIRCULATIONAHA.117.029652

7. Singh S, Sridhar P. A narrative review of sociodemographic risk and disparities in screening, diagnosis, treatment, and outcomes of the most common extrathoracic malignancies in the United States. J Thorac Dis. 2021;13(6):3827–3843. doi:10.21037/jtd-21-87

8. Agardh E, Allebeck P, Hallqvist J, Moradi T, Sidorchuk A. Type 2 diabetes incidence and socio-economic position: a systematic review and meta-analysis. Int J Epidemiol. 2011;40(3):804–818. doi:10.1093/ije/dyr029

9. Gupta A, Wilson LE, Pinheiro LC, et al. Association of educational attainment with cancer mortality in a national cohort study of black and white adults: A mediation analysis. SSM - Popul Health. 2023;24:101546. doi:10.1016/j.ssmph.2023.101546

10. Leung CW, Tester JM. The Association between Food Insecurity and Diet Quality Varies by Race/Ethnicity: An Analysis of National Health and Nutrition Examination Survey 2011-2014 Results. J Acad Nutr Diet. 2019;119(10):1676–1686. doi:10.1016/j.jand.2018.10.011

11. Leung CW, Wolfson JA, Brandt EJ, Rimm EB. Disparities in Cardiovascular Health by Food Security Status and Supplemental Nutrition Assistance Program Participation Using Life’s Essential 8 Metrics. JAMA Netw Open. 2023;6(6):e2321375. doi:10.1001/jamanetworkopen.2023.21375

12. Johnson AE, Herbert BM, Stokes N, Brooks MM, Needham BL, Magnani JW. Educational Attainment, Race, and Ethnicity as Predictors for Ideal Cardiovascular Health: From the National Health and Nutrition Examination Survey. J Am Heart Assoc Cardiovasc Cerebrovasc Dis. 2022;11(2):e023438. doi:10.1161/JAHA.121.023438

13. United States Department of Agriculture. Food Insecurity in the U.S. - Measurement. Accessed November 26, 2023. https://www.ers.usda.gov/topics/food-nutrition-assistance/food-security-in-the-u-s/measurement/

14. Liu J, Yi SS, Russo RG, et al. Trends and disparities in prevalence of cardiometabolic diseases by food security status in the United States. Nutr J. 2024;23:4. doi:10.1186/s12937-023-00910-4

15. Leung CW, Epel ES, Ritchie LD, Crawford PB, Laraia BA. Food insecurity is inversely associated with diet quality of lower-income adults. J Acad Nutr Diet. 2014;114(12):1943–1953.e2. doi:10.1016/j.jand.2014.06.353

16. Laraia BA. Food Insecurity and Chronic Disease. Adv Nutr. 2013;4(2):203–212. doi:10.3945/an.112.003277

17. National Research Council. Food Insecurity and Hunger in the United States: An Assessment of the Measure. The National Academies Press; 2006. 10.17226/11578.

18. U.S. Department of Agriculture, U.S. Department of Health and Human Services. Dietary Guidelines for Americans, 2020-2025. DietaryGuidelines.gov

19. Scientific Report of the 2025 Dietary Guidelines Advisory Committee: Advisory Report to the Secretary of Health and Human Services and Secretary of Agriculture. HHS and USDA; 2024. doi:10.52570/DGAC2025

20. Herman PM, Nguyen P, Sturm R. Diet quality improvement and 30-year population health and economic outcomes: a microsimulation study. Public Health Nutr. 25(5):1265–1273. doi:10.1017/S136898002100015X

21. Jardim TV, Mozaffarian D, Abrahams-Gessel S, et al. Cardiometabolic disease costs associated with suboptimal diet in the United States: A cost analysis based on a microsimulation model. Basu S, ed. PLOS Med. 2019;16(12):e1002981. doi:10.1371/journal.pmed.1002981

22. Thorndike AN, Gardner CD, Kendrick KB, et al. Strengthening US Food Policies and Programs to Promote Equity in Nutrition Security: A Policy Statement From the American Heart Association. Circulation. 2022;145(24):e1077–e1093. doi:10.1161/CIR.0000000000001072

23. Booker C, Rubio M. SNAP Nutrition Security Act.; 2023. https://www.booker.senate.gov/imo/media/doc/snap_nutrition_security_act_of_2023.pdf

24. ODPHP. White House Conference on Hunger, Nutrition and Health. Accessed May 20, 2024. https://health.gov/our-work/nutrition-physical-activity/white-house-conference-hunger-nutrition-and-health

25. Thompson G. *Farm, Food*, *and National Security Act of* 2024.; 2024. https://www.congress.gov/bill/118th-congress/house-bill/8467

26. Liu J, Micha R, Li Y, Mozaffarian D. Trends in Food Sources and Diet Quality Among US Children and Adults, 2003-2018. JAMA Netw Open. 2021;4(4):e215262. doi:10.1001/jamanetworkopen.2021.5262

27. Tao MH, Liu JL, Nguyen USDT. Trends in Diet Quality by Race/Ethnicity among Adults in the United States for 2011–2018. Nutrients. 2022;14(19):4178. doi:10.3390/nu14194178

28. Shan Z, Rehm CD, Rogers G, et al. Trends in Dietary Carbohydrate, Protein, and Fat Intake and Diet Quality Among US Adults, 1999-2016. JAMA. 2019;322(12):1178–1187. doi:10.1001/jama.2019.13771

29. Hu EA, Steffen LM, Coresh J, Appel LJ, Rebholz CM. Adherence to the Healthy Eating Index–2015 and Other Dietary Patterns May Reduce Risk of Cardiovascular Disease, Cardiovascular Mortality, and All-Cause Mortality. J Nutr. 2020;150(2):312–321. doi:10.1093/jn/nxz218

30. Zhang Y, Lu C, Li X, et al. Healthy Eating Index-2015 and Predicted 10-Year Cardiovascular Disease Risk, as Well as Heart Age. Front Nutr. 2022;9:888966. doi:10.3389/fnut.2022.888966

31. Blanton CA, Moshfegh AJ, Baer DJ, Kretsch MJ. The USDA Automated Multiple-Pass Method accurately estimates group total energy and nutrient intake. J Nutr. 2006;136(10):2594–2599. doi:10.1093/jn/136.10.2594

32. Ahluwalia N, Dwyer J, Terry A, Moshfegh A, Johnson C. Update on NHANES Dietary Data: Focus on Collection, Release, Analytical Considerations, and Uses to Inform Public Policy12. Adv Nutr. 2016;7(1):121–134. doi:10.3945/an.115.009258

33. Usual Dietary Intakes: Details of the Method | EGRP/DCCPS/NCI/NIH. Accessed December 19, 2023. https://epi.grants.cancer.gov/diet/usualintakes/details.html

34. Wingrove K, Lawrence MA, McNaughton SA. A Systematic Review of the Methods Used to Assess and Report Dietary Patterns. Front Nutr. 2022;9:892351. doi:10.3389/fnut.2022.892351

35. Schwingshackl L, Bogensberger B, Hoffmann G. Diet Quality as Assessed by the Healthy Eating Index, Alternate Healthy Eating Index, Dietary Approaches to Stop Hypertension Score, and Health Outcomes: An Updated Systematic Review and Meta-Analysis of Cohort Studies. J Acad Nutr Diet. 2018;118(1):74–100.e11. doi:10.1016/j.jand.2017.08.024

36. Krebs-Smith SM, Pannucci TE, Subar AF, et al. Update of the Healthy Eating Index: HEI-2015. J Acad Nutr Diet. 2018;118(9):1591–1602. doi:10.1016/j.jand.2018.05.021

37. Liese AD, Krebs-Smith SM, Subar AF, et al. The Dietary Patterns Methods Project: Synthesis of Findings across Cohorts and Relevance to Dietary Guidance. J Nutr. 2015;145(3):393–402. doi:10.3945/jn.114.205336

38. Panizza CE, Shvetsov YB, Harmon BE, et al. Testing the Predictive Validity of the Healthy Eating Index-2015 in the Multiethnic Cohort: Is the Score Associated with a Reduced Risk of All-Cause and Cause-Specific Mortality? Nutrients. 2018;10(4):452. doi:10.3390/nu10040452

39. Monteiro CA, Cannon G, Levy RB, et al. Ultra-processed foods: what they are and how to identify them. Public Health Nutr. 2019;22(5):936–941. doi:10.1017/S1368980018003762

40. Steele EM, O’Connor LE, Juul F, et al. Identifying and Estimating Ultraprocessed Food Intake in the US NHANES According to the Nova Classification System of Food Processing. J Nutr. 2023;153(1):225–241. doi:10.1016/j.tjnut.2022.09.001

41. National Health and Examination Survey. Anthropometry Procedures Manual.; 2017.

42. CDC. Defining Adult Overweight and Obesity. Centers for Disease Control and Prevention. June 3, 2022. Accessed January 27, 2024. https://www.cdc.gov/obesity/basics/adult-defining.html

43. National Insitutes of Health, National Heart, Lung, and Blood Institute. Clinical guidelines on the identification, evaluation, and treatment of overweight and obesity in adults: The evidence report. Obes Res. 1998;6(S2):S51–S210. doi:10.1002/j.1550-8528.1998.tb00690.x

44. 2018 AHA/ACC/AACVPR/AAPA/ABC/ACPM/ADA/AGS/APhA/ASPC/NLA/PCNA Guideline on the Management of Blood Cholesterol: A Report of the American College of Cardiology/American Heart Association Task Force on Clinical Practice Guidelines. doi:10.1161/CIR.0000000000000625

45. Miller M, Stone NJ, Ballantyne C, et al. Triglycerides and Cardiovascular Disease. Circulation. 2011;123(20):2292–2333. doi:10.1161/CIR.0b013e3182160726

46. American Diabetes Association. *Standards of Care in Diabetes—*2023 Abridged for Primary Care Providers. Clin Diabetes. 2023;41(1):4–31. doi:10.2337/cd23-as01

47. McCullough ML, Chantaprasopsuk S, Islami F, et al. Association of Socioeconomic and Geographic Factors With Diet Quality in US Adults. JAMA Netw Open. 2022;5(6):e2216406. doi:10.1001/jamanetworkopen.2022.16406

48. Liu J, Mozaffarian D. Trends in Diet Quality Among U.S. Adults From 1999 to 2020 by Race, Ethnicity, and Socioeconomic Disadvantage. Ann Intern Med. 2024;177(7):841–850. doi:10.7326/M24-0190

49. Baraldi LG, Martinez Steele E, Canella DS, Monteiro CA. Consumption of ultra-processed foods and associated sociodemographic factors in the USA between 2007 and 2012: evidence from a nationally representative cross-sectional study. BMJ Open. 2018;8(3):e020574. doi:10.1136/bmjopen-2017-020574

50. Bickel G, Nord M, Price C, Hamilton W, Cook J. Guide to Measuring Household Food Security, Revised 2000. U.S. Department of Agriculture, Food and Nutrition Service; 2000.

51. Gregory CA, Coleman-Jensen A. Food Insecurity, Chronic Disease, and Health Among Working-Age Adults. U.S. Department of Agriculture, Economic Research Service; 2017.

52. Lee EK, Donley G, Ciesielski TH, et al. Health outcomes in redlined versus non-redlined neighborhoods: A systematic review and meta-analysis. Soc Sci Med. 2022;294:114696. doi:10.1016/j.socscimed.2021.114696

53. Madlala SS, Hill J, Kunneke E, Lopes T, Faber M. Adult food choices in association with the local retail food environment and food access in resource-poor communities: a scoping review. BMC Public Health. 2023;23(1):1083. doi:10.1186/s12889-023-15996-y

54. Seligman HK, Levi R, Adebiyi VO, Coleman-Jensen A, Guthrie JF, Frongillo EA. Assessing and Monitoring Nutrition Security to Promote Healthy Dietary Intake and Outcomes in the United States. Annu Rev Nutr. 2023;43(1):409–429. doi:10.1146/annurev-nutr-062222-023359

