## Supplemental Tables and Figures for "Understanding Sociodemographic and Dietary Determinants of Cardiometabolic Risk: Cross-Sectional Evidence from the U.S. Healthy Eating Index to Inform Diet Quality Categories"

**Supplemental Figure 1: NHANES analytic flow-chart**

Analytic sample for adiposity and cardiometabolic outcomes

BMI = 21,955 (n = 213 missing)

Waist circumference = 21,395 (n = 773 missing)

HDL cholesterol = 21,148 (n = 1,020 missing)

LDL cholesterol = 10,020 (n = 279 missing)*

Triglycerides = 10,165 (n = 121 missing)*

Fasting glucose = 10,391*

*= fasting sub-sample

Did not complete two NHANES dietary assessments or implausible intake

n = 6,667

All adults ≥ 20 years in NHANES 2009-2018

n = 28,835

Analytic sample for sociodemographic and dietary outcomes

n = 22,168

**Supplemental Figure 2: Distributions of A) observed and B) usual HEI-2020 scores among US adults (≥20 years), 2009-2018 National Health and Nutrition Examination Surveys**
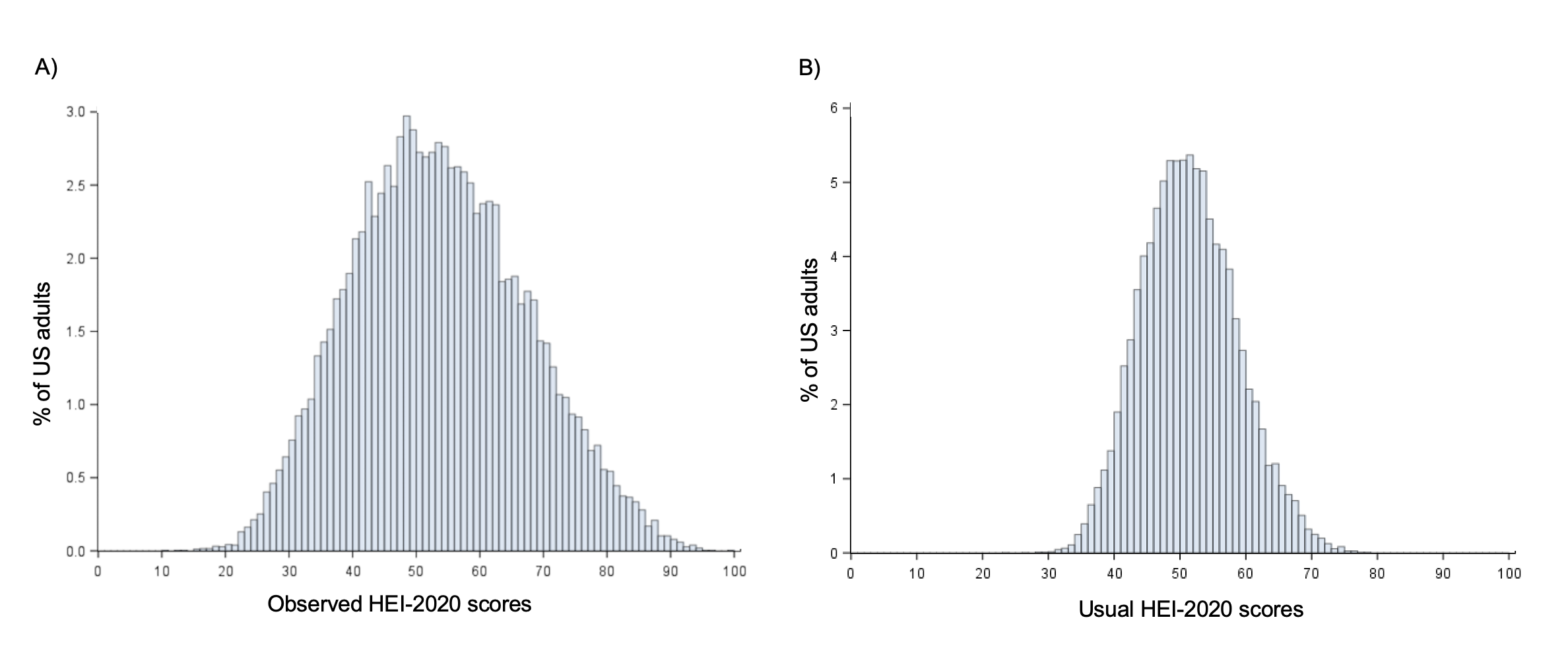

**Supplemental Figure 3: Proportion of diet quality categories across two-year cycles of NHANES, 2009-2018 National Health and Nutrition Examination Surveys**

|  | Very low diet quality | Low diet quality | Marginal diet quality | High diet quality |
| --- | --- | --- | --- | --- |
| 2017-18 | 43.5345 | 27.3023 | 18.4855 | 10.6777 |
| 2015-16 | 41.3578 | 25.8286 | 19.8076 | 13.0059 |
| 2013-14 | 41.1101 | 24.4877 | 21.0906 | 13.3117 |
| 2011-12 | 37.0814 | 27.7508 | 21.4877 | 13.6801 |
| 2009-10 | 38.9614 | 29.142 | 18.7358 | 13.1608 |

**Supplemental Table 1: Observational studies with significant associations between Healthy Eating Index scores and health outcomes**

| **Study** | **Population** | **Outcome** | **HEI version** | **Bottom quintile ranges** | **Top quintile ranges** |
| --- | --- | --- | --- | --- | --- |
| Kappeler, 2013^1^ | NHANES III (Men)^a^ | Total and cause-specific mortality | 2000 | < 51 | >80 |
| Kappeler, 2013^1^ | NHANES III (Women)^a^ | Total and cause-specific mortality | 2000 | < 51 | >80 |
| Reedy, 2014^2^ | NIH-AARP (Men) | Total and cause-specific mortality | 2010 | 18.2-55.2 | 74.1-96.1 |
| Reedy, 2014^2^ | NIH-AARP (Women) | Total and cause-specific mortality | 2010 | 18.5-59.3 | 76.4-96.2 |
| Yu, 2015^3^ | SCCS (Men)^b^ | Total and cause-specific mortality | 2010 | 13.7-45.3 | 64.8-95.2 |
| Yu, 2015^3^ | SCCS (Women)^b^ | Total and cause-specific mortality | 2010 | 16.9-48.8 | 70.2-96.7 |
| George, 2014^4^ | WHI^c^ | All-cause mortality; cardiovascular disease mortality; cancer mortality | 2010 | 18-58 | 76-95 |
| Harmon, 2015^5^ | MEC (Men)^d^ | All-cause mortality; cardiovascular disease mortality; cancer mortality | 2010 | 13.4-55.9 | 74.8-99.9 |
| Harmon, 2015^5^ | MEC (Women)^d^ | All-cause mortality; cardiovascular disease mortality; cancer mortality | 2010 | 21.2-60.3 | 79.0-100 |
| Panizza, 2018^6^ | MEC (Men)^d^ | All-cause mortality; cardiovascular disease mortality; cancer mortality | 2015 | 17-9-56.1 | 74.1-98.7 |
| Panizza, 2018^6^ | MEC (Women)^d^ | All-cause mortality; cardiovascular disease mortality; cancer mortality | 2015 | 23.5-59.8 | 78.1-99.8 |
| Deshmukh, 2018^7^ | NHANES^a^ | All-cause mortality and cancer mortality among cancer survivors | 1994-1996 | ≤57.5 | ≥77 |
| George, 2011^8^ | HEAL^e^ | All-cause mortality; breast cancer mortality | 2005 | 50.1 (mean) | 79.0 (mean) |
| Hu, 2020^9^ | ARIC^f^ | All-cause mortality; cardiovascular disease incidence; cardiovascular disease mortality | 2015 | 60 (median) | 81 (median) |
| Xu, 2020^10^ | ARIC^f^ | Cardiovascular disease and type 2 diabetes | 2015 | 58.2 (mean) | 82.5 (mean) |
| McCullough, 2000a^11^ | NHS^g^ | Cardiovascular disease, cancer, or death | 2000 | 48.8 | 82.4 |
| McCullough, 2000b^12^ | HPFS^h^ | Cardiovascular disease, cancer, or death | 2000 | 21.6-58.2 | 78.7-97.4 |
| Park, 2017^13^ | MEC^d^ | Colorectal cancer | 2010 | 21.1 - 62.4 | 79.5-98.8 |
| Pelser, 2014^14^ | NIH-AARP^b^ | Colorectal cancer | 2005 | 21.8-56.7 | 77.0-90.2 |
| Reedy, 2008^15^ | NIH-AARP (Men) | Colorectal cancer | 2005 | 21-55 | 76-97 |
| Reedy, 2008^15^ | NIH-AARP (Women) | Colorectal cancer | 2005 | 20-60 | 79-94 |
| Thomson, 2014^16^ | WHI^c^ | Colorectal cancer | 2010 | 15.65-46.09 | 61.51-94.43 |
| Vargas, 2016^17^ | WHI^c^ | Colorectal cancer | 2010 | 15.65-46.09 | 61.51-94.43 |
| George, 2015^18^ | WHI^c^ | Endometrial cancer | 2010 | 17-56 | 76-95 |
| Li, 2013^19^ | NIH-AARP | Esophageal cancer | 2005 | 50 (median) | 80 (median) |
| Li, 2014a^20^ | NIH-AARP (Men) | Head and neck cancer | 2005 | 49 (median) | 79 (median) |
| Li, 2014^20(p20)^ | NIH-AARP (Women) | Head and neck cancer | 2005 | 54 (median) | 81 (median) |
| Bogumil, 2019^21^ | MEC^d^ | Hepatocellular carcinoma and chronic liver disease | 2010 | 13.5-57.4 | 76.8-99.9 |
| Li, 2014b^22^ | NIH-AARP | Liver cancer and liver disease mortality | 2019 | 52 (median) | 79 (median) |
| Anic, 2016^23^ | NIH-AARP (Men) | Lung cancer | 2010 | 20.7-56.8 | 77.0-96.6 |
| Anic, 2016^23(p2)^ | NIH-AARP (Women) | Lung cancer | 2010 | 19.9-56.8 | 77.0-94.0 |
| Park, 2021^24^ | MEC^d^ | Lung cancer | 2010 | 17.9-58.2 | 76.7-100 |
| Chiuve, 2012^25^ | NHS^g^ | Major chronic disease incidence (coronary heart disease, stroke, angina, diabetes, cancer) | 2005 | < 53.5 | > 71.3 |
| Chiuve, 2012^25^ | HPFS^h^ | Major chronic disease incidence (coronary heart disease, stroke, angina, diabetes, cancer) | 2005 | < 53.6 | > 73.2 |
| Haring, 2016^26^ | WHI^c^ | Mild cognitive impairment or probable dementia | 2010 | < 51 | >71.3 |
| Xie, 2014^27^ | NHS^g^ | Ovarian cancer | 2005 | < 56 | >74 |
| Arem, 2013^28^ | NIH-AARP (Men) | Pancreatic cancer | 2005 | 20.6-54.7^i^ | 75.6-96.7^i^ |
| Arem, 2013^28^ | NIH-AARP (Women) | Pancreatic cancer | 2005 | 19.7-60.4^i^ | 78.6-94.0^i^ |
| Bosire, 2013^29^ | NIH-AARP (Men) | Prostate cancer | 2005 | 21-55^j^ | 76-97^j^ |
| Cespedes, 2016^30^ | WHI^c^ | Type 2 diabetes | 2010 | 18-57 | 77-95 |
| de Koning, 2011^31^ | HPFS^h^ | Type 2 diabetes | 2005 | 24-59 | 77-99 |
| Jacobs, 2015^32^ | MEC (Men)^d^ | Type 2 diabetes | 2010 | 21-57 | 76-100 |
| Jacobs, 2015^32^ | MEC (Women)^d^ | Type 2 diabetes | 2010 | 20-61 | 80-100 |

^a^ National Health and Nutrition Examination Survey

^b^ Southern Community Cohort Study

^c^ Women’s Health Initiative

^d^ Multiethnic Cohort

^e^ Health, Eating, Activity, and Lifestyle Study

^f^ Atherosclerosis Risk in Communities Study

^g^ Nurses’ Health Study

^h^ Health Professionals Follow-Up Study

^i^ Quartiles used

^j^ One standard deviation above and below the mean

**Supplemental Table 2: Multivariate-adjusted usual intakes of total energy, Nova food groups, foods, and nutrients of interest stratified by diet quality categories (usual HEI-2020 scores), NHANES 2009-2018**

|  | **Diet quality category** | | | | ***P-trend*** |
| --- | --- | --- | --- | --- | --- |
|  | **Very low^a^**  **Mean** | **Low^a^**  **Mean** | **Marginal^a^**  **Mean** | **High^a^**  **Mean** |  |
| Total energy (kcal/day) | 2146 | 2149 | 2147 | 2146 | *0.04* |
| Unprocessed/ minimally processed foods (% energy/day) | 29.0^b^ | 32.6^b^ | 35.7^b^ | 37.9 | *<0.01* |
| Ultra-processed foods (% energy/day) | 58.1^b^ | 53.0^b^ | 49.3^b^ | 47.1 | *<0.01* |
| Foods (cup or oz eq per 1000 kcal) |  |  |  |  |  |
| Fruit and vegetables, excluding juice and white potatoes (cup eq/day) | 0.8^b^ | 1.2^b^ | 1.6^b^ | 1.8 | *<0.01* |
| Refined grains (oz eq/day) | 3.0^b^ | 2.7^b^ | 2.4^b^ | 2.2 | *<0.01* |
| Whole grains (oz eq/day) | 0.5^b^ | 0.6^b^ | 0.7^b^ | 0.9 | *<0.01* |
| Red and processed meat (oz eq/day) | 1.4^b^ | 1.3^b^ | 1.3^b^ | 1.2 | *<0.01* |
| Seafood (oz eq/day) | 1.0^b^ | 1.1^b^ | 1.3^b^ | 1.6 | *<0.01* |
| Nutrients per 1,000 calories |  |  |  |  |  |
| Saturated fat (% energy/day) | 11.3^b^ | 10.6^b^ | 9.9^b^ | 9.2 | *<0.01* |
| Dietary fiber (g/day) | 7.4^b^ | 9.2^b^ | 11.4^b^ | 13.8 | *<0.01* |
| Sodium (mg/day) | 1734^b^ | 1707^b^ | 1671^b^ | 1569 | *<0.01* |
| Potassium (mg/day) | 1237^b^ | 1340^b^ | 1437^b^ | 1514 | *<0.01* |
| Calcium (mg/day) | 462^b^ | 485^b^ | 509^b^ | 544 | *<0.01* |
| Added sugar (% energy/day) | 14.5^b^ | 11.8^b^ | 10.5 | 10 | *<0.01* |

^a^ Values adjusted for age, sex, race and ethnicity, birthplace, educational attainment, employment status, marital status, household income, and household food security

^b^ Significant difference compared to the high diet quality category

**Supplemental Table 3: Multivariate-adjusted adiposity and cardiometabolic markers stratified by diet quality categories (usual HEI-2020 scores), NHANES 2009-2018**

|  | **Diet Quality category** | | | | ***P-trend*** |
| --- | --- | --- | --- | --- | --- |
|  | **Very Low^a^**  **Mean** | **Low^a^**  **Mean** | **Marginal^a^**  **Mean** | **High^a^**  **Mean** |  |
| BMI (kg/m2) | 29.9^b^ | 28.7^b^ | 27.3^b^ | 25 | *<0.01* |
| *% with obesity* | *46.1*^b^ | *37.6*^b^ | *26.6* | *11.7* | *<0.01* |
| Waist circumference (cm) | 100.4^b^ | 97.5^b^ | 93.9^b^ | 87.8 | *<0.01* |
| *% with high WC* | *60.7*^b^ | *58.8*^b^ | *51.1* | *31.7* | *<0.01* |
| HDL cholesterol (mg/dL) | 50.6^b^ | 53.8^b^ | 56.6 | 61.7 | *<0.01* |
| *% <40 mg/dL* | *25.3*^b^ | *16.2* | *13.0* | *8.9* | *<0.01* |
| LDL cholesterol (mg/dL) | 112.6 | 110.9 | 108.9 | 106.8 | *0.01* |
| *% ≥100 mg/dL* | *62.0* | *64.6* | *63.4* | *62.1* | *0.09* |
| Triglycerides (mg/dL) | 122.8^b^ | 118.3^b^ | 106.6 | 96.8 | *<0.01* |
| *% ≥150 mg/dL* | *25.8*^b^ | *24.3*^b^ | *19.2* | *13.4* | *<0.01* |
| Fasting glucose (mg/dL) | 107.1^b^ | 105.9^b^ | 101.6 | 98.7 | *<0.01* |
| % >100 mg/dL | *46.7*^b^ | *47.6* | *44.4* | *41.9* | *0.01* |

^a^ Values adjusted for age, sex, race and ethnicity, birthplace, educational attainment, employment status, marital status, household income, and household food security

^b^ Significant difference compared to the high diet quality category

**References**

1.	Kappeler R, Eichholzer M, Rohrmann S. Meat consumption and diet quality and mortality in NHANES III. *Eur J Clin Nutr*. 2013;67(6):598-606. doi:10.1038/ejcn.2013.59

2.	Reedy J, Krebs-Smith SM, Miller PE, et al. Higher Diet Quality Is Associated with Decreased Risk of All-Cause, Cardiovascular Disease, and Cancer Mortality among Older Adults. *J Nutr*. 2014;144(6):881-889. doi:10.3945/jn.113.189407

3.	Yu D, Sonderman J, Buchowski MS, et al. Healthy Eating and Risks of Total and Cause-Specific Death among Low-Income Populations of African-Americans and Other Adults in the Southeastern United States: A Prospective Cohort Study. Stuckler D, ed. *PLOS Med*. 2015;12(5):e1001830. doi:10.1371/journal.pmed.1001830

4.	George SM, Ballard-Barbash R, Manson JE, et al. Comparing Indices of Diet Quality With Chronic Disease Mortality Risk in Postmenopausal Women in the Women’s Health Initiative Observational Study: Evidence to Inform National Dietary Guidance. *Am J Epidemiol*. 2014;180(6):616-625. doi:10.1093/aje/kwu173

5.	Harmon BE, Boushey CJ, Shvetsov YB, et al. Associations of key diet-quality indexes with mortality in the Multiethnic Cohort: the Dietary Patterns Methods Project. *Am J Clin Nutr*. 2015;101(3):587-597. doi:10.3945/ajcn.114.090688

6.	Panizza CE, Shvetsov YB, Harmon BE, et al. Testing the Predictive Validity of the Healthy Eating Index-2015 in the Multiethnic Cohort: Is the Score Associated with a Reduced Risk of All-Cause and Cause-Specific Mortality? *Nutrients*. 2018;10(4):452. doi:10.3390/nu10040452

7.	Deshmukh AA, Shirvani SM, Likhacheva A, Chhatwal J, Chiao EY, Sonawane K. The Association Between Dietary Quality and Overall and Cancer-Specific Mortality Among Cancer Survivors, NHANES III. *JNCI Cancer Spectr*. 2018;2(2):pky022. doi:10.1093/jncics/pky022

8.	George SM, Irwin ML, Smith AW, et al. Postdiagnosis diet quality, the combination of diet quality and recreational physical activity, and prognosis after early-stage breast cancer. *Cancer Causes Control*. 2011;22(4):589-598. doi:10.1007/s10552-011-9732-9

9.	Hu EA, Steffen LM, Coresh J, Appel LJ, Rebholz CM. Adherence to the Healthy Eating Index–2015 and Other Dietary Patterns May Reduce Risk of Cardiovascular Disease, Cardiovascular Mortality, and All-Cause Mortality. *J Nutr*. 2020;150(2):312-321. doi:10.1093/jn/nxz218

10. Xu Z, Steffen LM, Selvin E, Rebholz CM. Diet quality, change in diet quality and risk of incident CVD and diabetes. *Public Health Nutr*. 2020;23(2):329-338. doi:10.1017/S136898001900212X

11. McCullough ML, Feskanich D, Stampfer MJ, et al. Adherence to the Dietary Guidelines for Americans and risk of major chronic disease in women. *Am J Clin Nutr*. 2000;72(5):1214-1222. doi:10.1093/ajcn/72.5.1214

12. McCullough ML, Feskanich D, Rimm EB, et al. Adherence to the *Dietary Guidelines for Americans* and risk of major chronic disease in men. *Am J Clin Nutr*. 2000;72(5):1223-1231. doi:10.1093/ajcn/72.5.1223

13. Park SY, Boushey CJ, Wilkens LR, Haiman CA, Le Marchand L. High-Quality Diets Associate With Reduced Risk of Colorectal Cancer: Analyses of Diet Quality Indexes in the Multiethnic Cohort. *Gastroenterology*. 2017;153(2):386-394.e2. doi:10.1053/j.gastro.2017.04.004

14. Pelser C, Arem H, Pfeiffer RM, et al. Prediagnostic lifestyle factors and survival after colon and rectal cancer diagnosis in the National Institutes of Health (NIH)-AARP Diet and Health Study. *Cancer*. 2014;120(10):1540-1547. doi:10.1002/cncr.28573

15. Reedy J, Mitrou PN, Krebs-Smith SM, et al. Index-based Dietary Patterns and Risk of Colorectal Cancer: The NIH-AARP Diet and Health Study. *Am J Epidemiol*. 2008;168(1):38-48. doi:10.1093/aje/kwn097

16. Thomson CA, Crane TE, Wertheim BC, et al. Diet Quality and Survival After Ovarian Cancer: Results From the Women’s Health Initiative. *JNCI J Natl Cancer Inst*. 2014;106(11):dju314. doi:10.1093/jnci/dju314

17. Vargas AJ, Neuhouser ML, George SM, et al. Diet Quality and Colorectal Cancer Risk in the Women’s Health Initiative Observational Study. *Am J Epidemiol*. 2016;184(1):23-32. doi:10.1093/aje/kwv304

18. George SM, Ballard R, Shikany JM, Crane TE, Neuhouser ML. A prospective analysis of diet quality and endometrial cancer among 84,415 postmenopausal women in the Women’s Health Initiative. *Ann Epidemiol*. 2015;25(10):788-793. doi:10.1016/j.annepidem.2015.05.009

19. Li W, Park Y, Wu JW, et al. Index-based Dietary Patterns and Risk of Esophageal and Gastric Cancer in a Large Cohort Study. *Clin Gastroenterol Hepatol*. 2013;11(9):1130-1136.e2. doi:10.1016/j.cgh.2013.03.023

20. Li WQ, Park Y, Wu JW, et al. Index-based dietary patterns and risk of head and neck cancer in a large prospective study. *Am J Clin Nutr*. 2014;99(3):559-566. doi:10.3945/ajcn.113.073163

21. Bogumil D, Park SY, Le Marchand L, et al. High‐Quality Diets Are Associated With Reduced Risk of Hepatocellular Carcinoma and Chronic Liver Disease: The Multiethnic Cohort. *Hepatol Commun*. 2019;3(3):437. doi:10.1002/hep4.1313

22. Li WQ, Park Y, McGlynn KA, et al. Index-based dietary patterns and risk of incident hepatocellular carcinoma and mortality from chronic liver disease in a prospective study. *Hepatology*. 2014;60(2):588-597. doi:10.1002/hep.27160

23. Anic GM, Park Y, Subar AF, Schap TE, Reedy J. Index-based dietary patterns and risk of lung cancer in the NIH–AARP diet and health study. *Eur J Clin Nutr*. 2016;70(1):123-129. doi:10.1038/ejcn.2015.122

24. Park SY, Boushey CJ, Shvetsov YB, et al. Diet Quality and Risk of Lung Cancer in the Multiethnic Cohort Study. *Nutrients*. 2021;13(5):1614. doi:10.3390/nu13051614

25. Chiuve SE, Fung TT, Rimm EB, et al. Alternative Dietary Indices Both Strongly Predict Risk of Chronic Disease. *J Nutr*. 2012;142(6):1009-1018. doi:10.3945/jn.111.157222

26. Haring B, Wu C, Mossavar-Rahmani Y, et al. No Association between Dietary Patterns and Risk for Cognitive Decline in Older Women with 9-Year Follow-Up: Data from the Women’s Health Initiative Memory Study. *J Acad Nutr Diet*. 2016;116(6):921-930.e1. doi:10.1016/j.jand.2015.12.017

27. Xie J, Poole EM, Terry KL, et al. A prospective cohort study of dietary indices and incidence of epithelial ovarian cancer. *J Ovarian Res*. 2014;7:112. doi:10.1186/s13048-014-0112-4

28. Arem H, Reedy J, Sampson J, et al. The Healthy Eating Index 2005 and Risk for Pancreatic Cancer in the NIH–AARP Study. *JNCI J Natl Cancer Inst*. 2013;105(17):1298-1305. doi:10.1093/jnci/djt185

29. Bosire C, Stampfer MJ, Subar AF, et al. Index-based Dietary Patterns and the Risk of Prostate Cancer in the NIH-AARP Diet and Health Study. *Am J Epidemiol*. 2013;177(6):504-513. doi:10.1093/aje/kws261

30. Cespedes EM, Hu FB, Tinker L, et al. Multiple Healthful Dietary Patterns and Type 2 Diabetes in the Women’s Health Initiative. *Am J Epidemiol*. 2016;183(7):622-633. doi:10.1093/aje/kwv241

31. de Koning L, Chiuve SE, Fung TT, Willett WC, Rimm EB, Hu FB. Diet-Quality Scores and the Risk of Type 2 Diabetes in Men. *Diabetes Care*. 2011;34(5):1150-1156. doi:10.2337/dc10-2352

32. Jacobs S, Harmon BE, Boushey CJ, et al. A priori-defined diet quality indexes and risk of type 2 diabetes: the Multiethnic Cohort. *Diabetologia*. 2015;58(1):98-112. doi:10.1007/s00125-014-3404-8
